# COVID-19 vaccine effectiveness against severe disease from the Omicron BA.1 and BA.2 subvariants – surveillance results from southern Sweden, December 2021 to March 2022

**DOI:** 10.1101/2022.04.14.22273896

**Authors:** Jonas Björk, Carl Bonander, Mahnaz Moghaddassi, Magnus Rasmussen, Ulf Malmqvist, Malin Inghammar, Fredrik Kahn

## Abstract

We compared vaccine effectiveness (VE) against severe COVID-19 during calendar periods from December 2021 to March 2022 when Omicron BA.1 and BA.2, respectively, were the dominating virus variants in Scania county, Sweden. We used continuous density case-control sampling matched for sex and age, and with further adjustment for differences in comorbidities and prior infection. VE remained relatively stable after the transition from BA.1 to BA.2 among people with at least three doses but decreased markedly among those with only two doses. Protection from prior infection was also lower after the transition to BA.2. These findings suggest that booster vaccination is needed to maintain sufficient protection against severe COVID-19.

---

The severe acute respiratory syndrome coronavirus 2 (SARS-CoV-2) variant of concern (VOC) Omicron (Phylogenetic Assignment of Named Global Outbreak (Pango) lineage designation B.1.1.529) has two genetically divergent subvariants, BA.1 and BA.2. A comparison of secondary attack rates from Denmark suggests that the subvariant BA.2 carries a transmission advantage compared to BA.1 (1), which may explain why BA.2 has rapidly replaced BA.1 as the dominant subvariant in several countries. A study from UK suggests similar vaccine protection against symptomatic disease from BA.2 as for BA.1 (2). However, few studies have compared the protection against severe disease from these two Omicron subvariants. Recent findings from Qatar suggested lower vaccine protection against hospitalizations and deaths from BA.2 but with a very wide confidence interval (3).

The aim of the present study was to compare COVID-19 vaccine effectiveness against severe disease from the Omicron BA.1 and BA.2 subvariants. The study was conducted in Scania county, southern Sweden, one of the first regions in the world where BA.2 became dominant.

## Study design and data extraction

The study cohort included all persons residing in Scania county (Skåne), southern Sweden, on 27 December 2020 (baseline) when vaccinations started (n = 1,384,531) (4, 5) and were followed longitudinally for positive SARS-CoV-2 tests, hospitalisations and assessment of disease severity. Individuals who died or moved away from the region were censored on the date of death or relocation. In the present study we used data from routine sequencing of samples of infected cases in Scania county to compare vaccine effectiveness during three specific calendar periods (Supplementary Table S1): (i) Omicron BA.1 as the dominating VOC, 2021 week 52 and 2022 week 1 (BA.1: 60%, Delta: 25%, BA.2: 15%), (ii) transition period, 2022 week 2–3 (BA.1: 47%, BA.2: 49%, Delta: 4.5%), and (iii) Omicron BA.2 as the dominating VOC, 2022 week 4-11 (until date of data extraction 15 March; BA.2: 82%, BA. 17%:, Delta: 0.5%).

Data from national and regional register holders were linked using the personal identification number assigned to all Swedish residents. Weekly updates on vaccination date, type of vaccine and dose were obtained from the National Vaccination Register, and data on COVID-19 cases (defined by a positive SARS-CoV-2 test results) from the electronic system SMINet, both kept at the Public Health Agency of Sweden. Regional health registers were used as complementary data sources to provide data on positive tests rapidly, and to assess comorbidities and disease outcomes.

Comorbidities were defined from diagnoses in inpatient or specialised care at any time point during the 5 years before baseline in the following disease groups (see Supplementary Table S2 for a detailed list): cardiovascular diseases, diabetes or obesity, kidney or liver diseases, respiratory diseases, neurological diseases, cancer or immunosuppressed states, and other conditions and diseases (Down syndrome, HIV, sickle cell anaemia, drug addiction, thalassaemia or mental health disorder). The number of comorbidities in these groupings was counted and used in the analyses. A severe COVID-19 case was defined as at least 24 h hospitalisation 5 days before until 14 days after a positive test and with a need of oxygen supply ≥ 5 L/min or admittance to an intensive care unit (ICU).

## Vaccine effectiveness

We used continuous density case-control sampling (6) nested within the study cohort together with conditional logistic regression to estimate vaccine effectiveness (VE) against severe COVID-19. For each severe case, 10 controls without a positive test the same week as the case or 90 days prior were randomly selected from the underlying study cohort, matched with respect to sex and age (five-year groups). Only vaccine doses obtained more than 7 days before the case date were counted in the analyses. BNT16b2 mRNA (Comirnaty, Pfizer-BioNTech) was most frequently used vaccine type with 77% of all administrated doses in the study cohort.

A total of 593 severe COVID-cases occurred during follow up, corresponding to 65, 78 and 56 cases weekly during the Omicron BA.1, transition and Omicron BA.2 periods, respectively (Table 1). Severe cases during BA.2 were older and had a more even sex distribution than during BA.1. The overrepresentation of persons born abroad among severe cases observed during BA.1 was less marked during BA.2, but the presence of comorbidities was similar. The monthly surveillance of population protection against severe COVID-19 after at least two doses was high before the follow up of the present study started (median VE 89% during March – November 2021) and remained stable also during Delta and Omicron BA.1 dominance in December 2021 (Figure 1). The transition from Omicron BA.1 to BA.2 in January – February 2022 was associated with a decline in population protection.

**Table 1.**
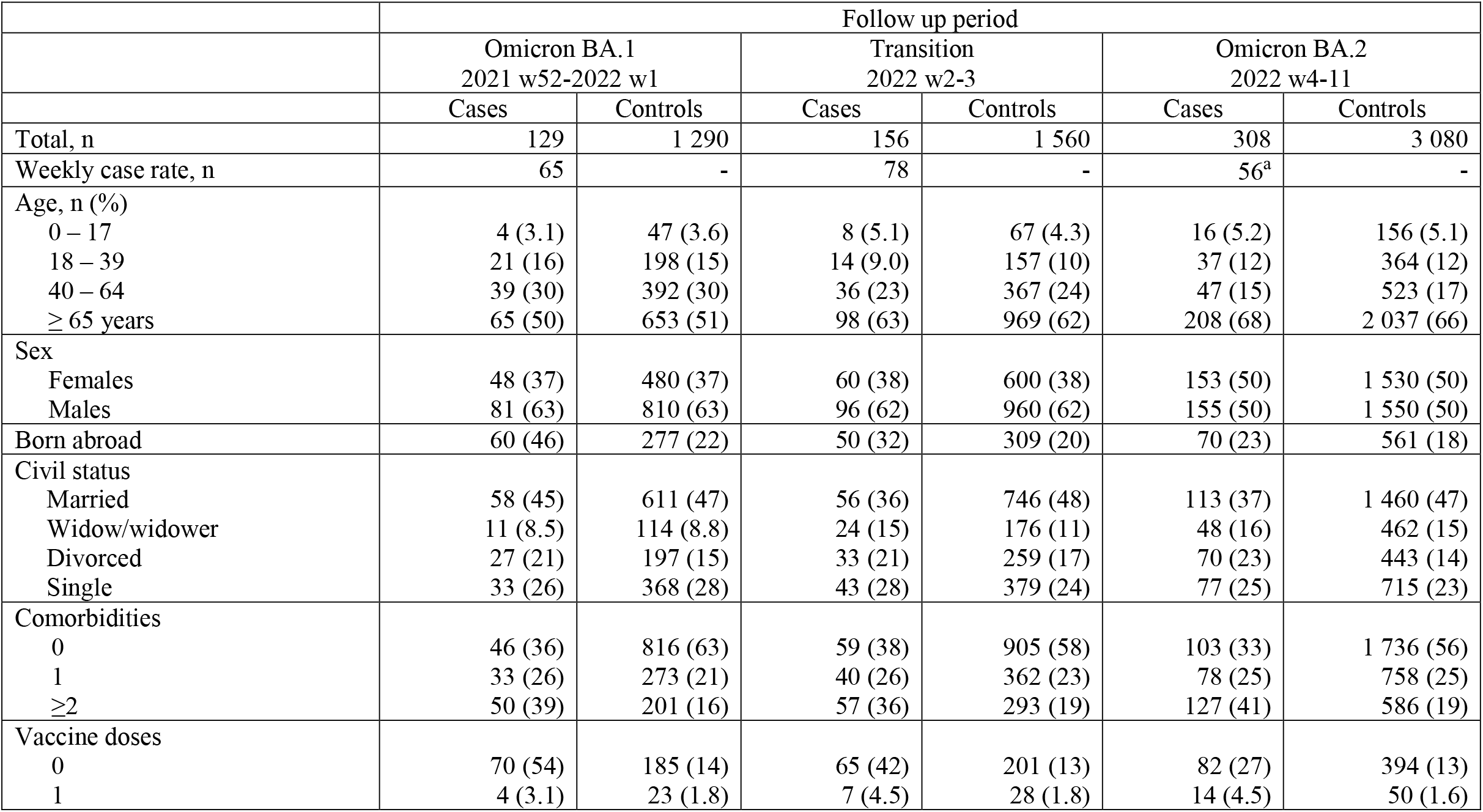
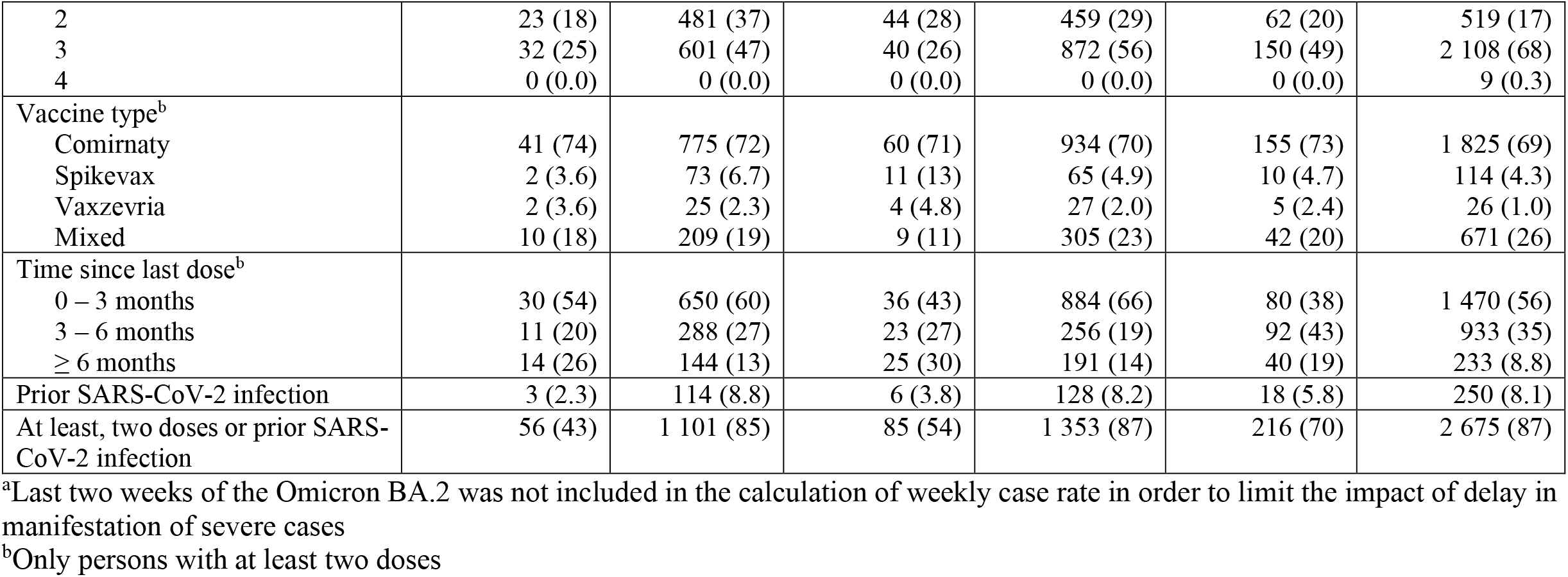
Characteristics of the severe COVID-19 cases (N = 593) and sex and age matched controls (N = 5 930), stratified by follow up period for monitoring of vaccine effectiveness.

**Figure 1.**
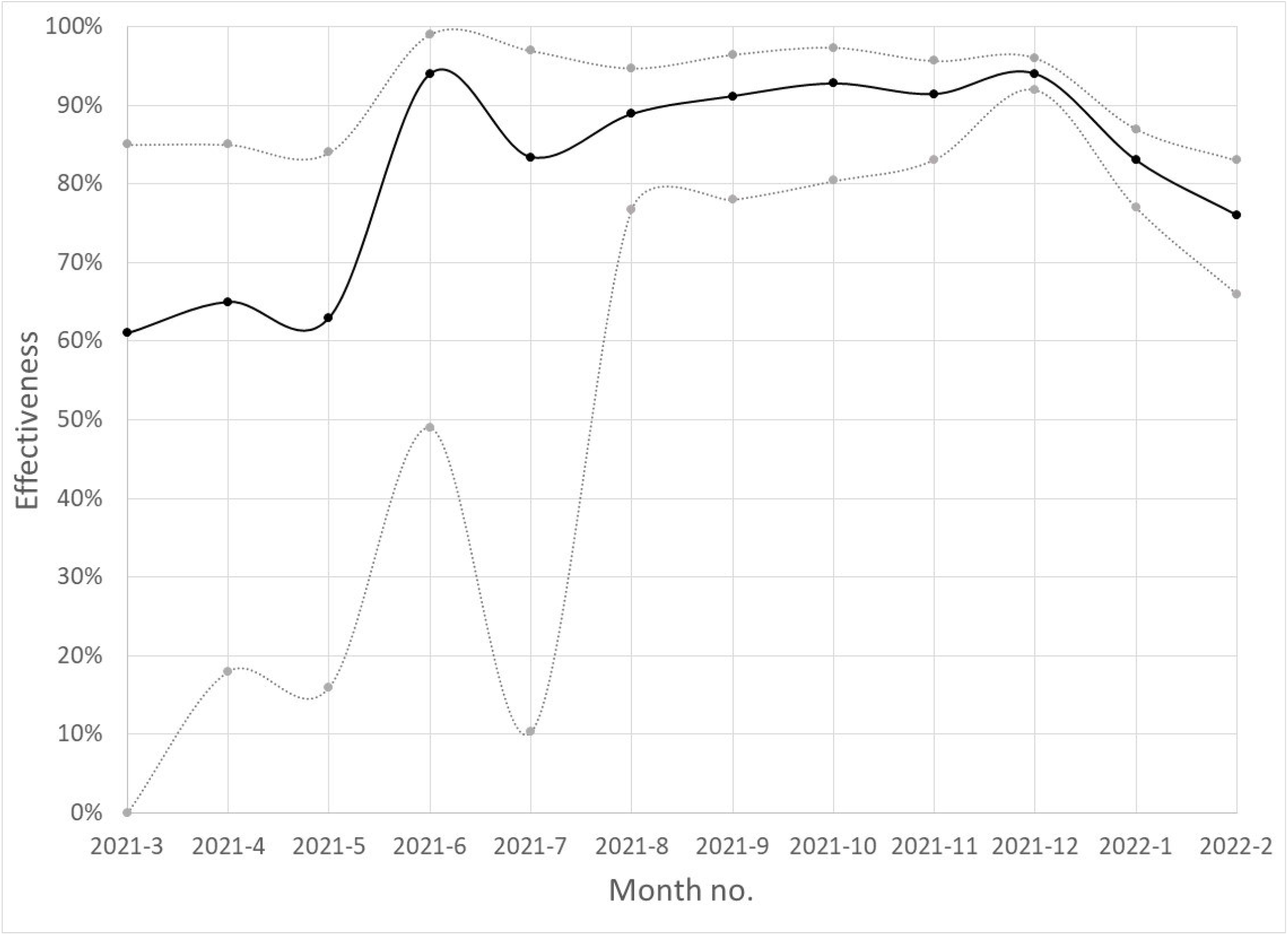
Surveillance in Scania county, southern Sweden, of population protection after at least two doses against severe COVID-19, monthly July 2021 - February 2022, (solid black curves). Estimated vaccine effectiveness obtained from conditional logistic regression for age and sex matched case and controls (1:10). Grey dotted lines represent 95% confidence intervals.

In more detailed analyses of the change in VE, number of vaccine doses were grouped as 0-1, 2 or at least 3 and included adjustment for the number of comorbidities and infection at least 90 days before the case date. Whereas the VE after at least three doses remained stable during follow up of the present study, VE after two doses declined substantially from 90% (95% confidence interval [CI]) 78 – 95%) during Omicron BA.1 dominance to 54% (95% CI 13 – 75%) during BA.2 dominance (Figure 2 and Supplementary Table 3). This decline was consistently noted across subgroups of age, sex and comorbidities.

**Figure 2.**
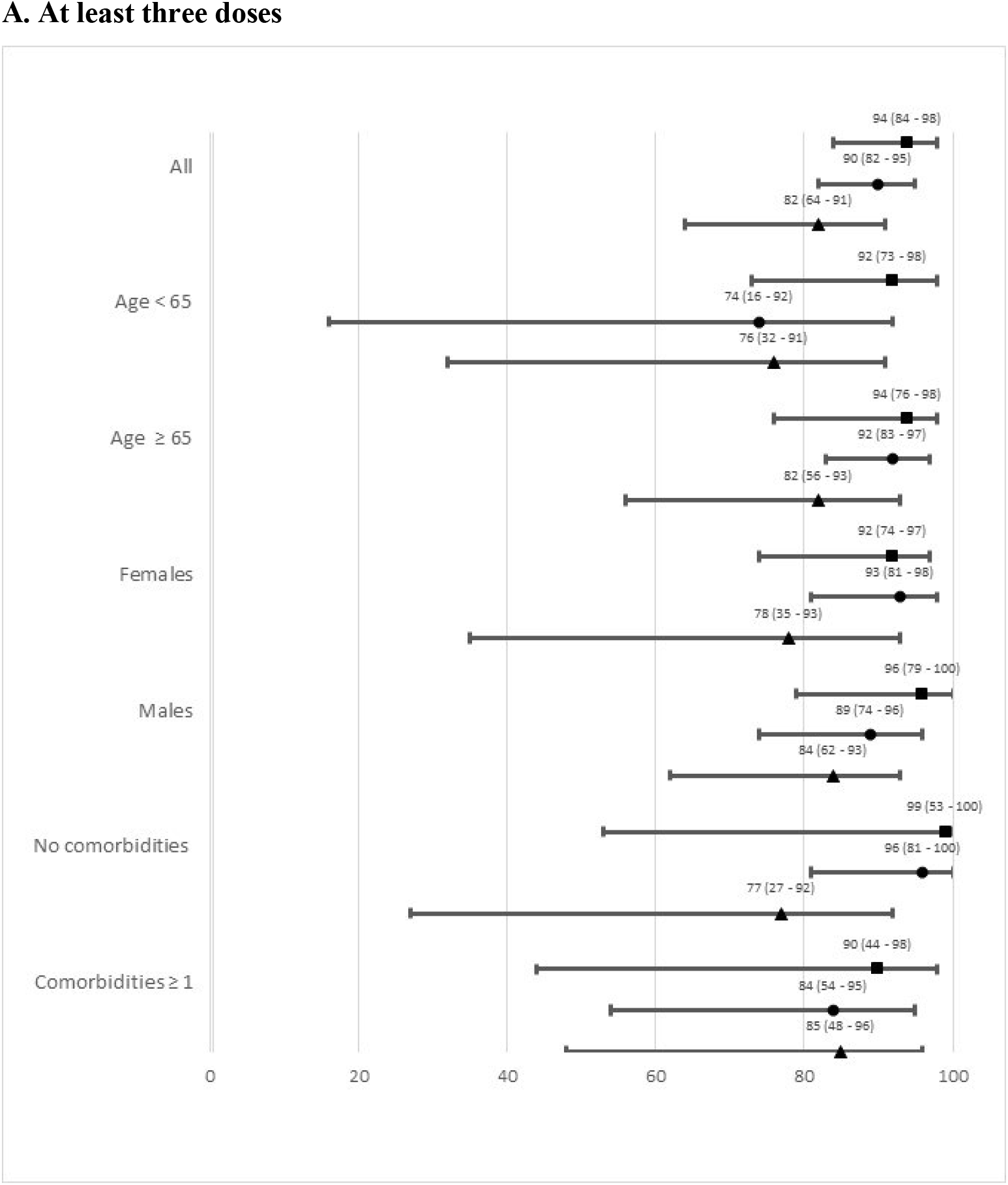
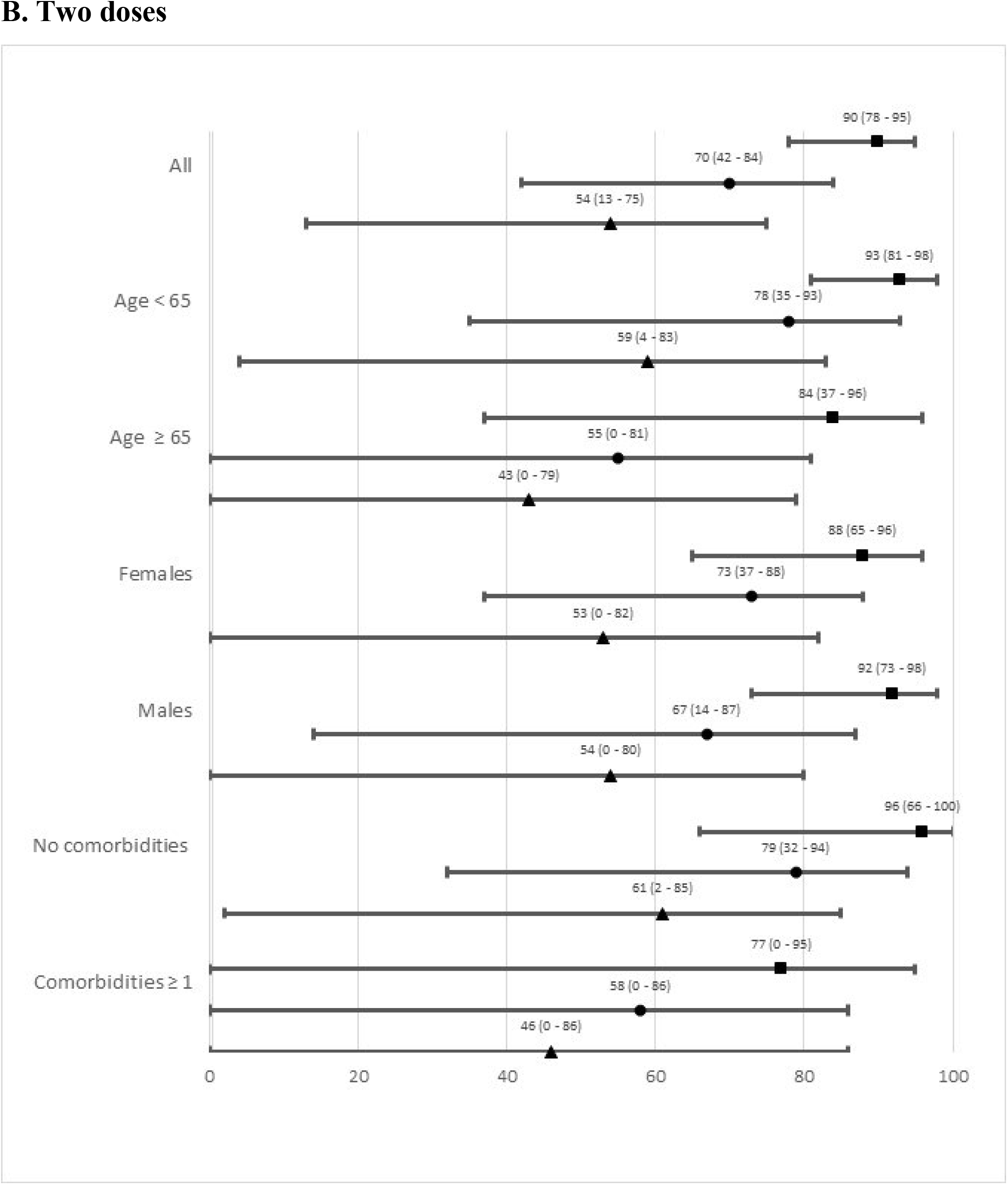
Vaccine effectiveness against severe COVID-19 in each follow up period (■ = Omicron BA.1, ● = Transition, ▲ = Omicron BA.2) after **A**. at least three or **B**. two doses. Estimates were obtained from conditional logistic regression for age and sex matched case and controls (1:10), and with adjustment for comorbidities (0, 1, ≥2) and infection at least 90 days prior the case date. Results are presented overall and stratified by age, sex and comorbidities.

In relative terms, surviving a prior SARS-CoV-2 infection offered stronger protection against new onset of severe COVID-19 among unvaccinated than among vaccinated (Table 2). However, this protection decreased markedly in both groups during the transition from Omicron BA.1 to BA.2.

**Table 2.** Protection against severe COVID-19 associated with prior SARS-CoV-2 infection at least 90 days before the case date in each follow up period. Estimates were obtained from unconditional logistic regression with adjustment for age (<18, 18 – 39, 40 – 64, ≥ 65 years), sex and comorbidities (0, 1, ≥ 2). Results are presented overall and stratified for vaccination status.

|  | Follow up period |  |  |
| --- | --- | --- | --- |
|  | Omicron BA.1<br>2021 w52-2022 w1 | Transition<br>2022 w2-3 | Omicron BA.2<br>2022 w4-11 |
|  | Protection (95% CI)<sup>a</sup> | Protection (95% CI)<sup>a</sup> | Protection (95% CI)<sup>a</sup> |
| All | 80 (36 – 94) | 62 (13 – 83) | 38 (0 – 61) |
| Vaccination status |  |  |  |
| 0 – 1 dose | 90 (28 – 99) | 87 (8 – 98) | 59 (0 – 85) |
| ≥ 2 doses | 61 (0 – 90) | 37 (0 – 75) | 23 (0 – 55) |

<sup>a</sup>Protection was estimated as $100 \times (1 - \text{OR})$ . OR = odds ratio, CI = confidence interval.

## Ethical statement

Ethical approval was obtained from the Swedish Ethical Review Authority (2021-00059).

## Discussion

A salient finding of the present study was the marked decline in protection against severe COVID-19 during Omicron BA.2 dominance among persons with two vaccine doses only. The decline occurred quite rapidly and thus cannot be explained by waning VE alone. A more likely explanation is increased immune evasiveness properties of BA.2, which gives this genetically subvariant a competitive advantage in highly vaccinated populations (1). The relatively stable protection among individuals with at least three doses during follow up in our study suggests that a very robust immune response is necessary to confer protection also against BA.2.

Although the overall vaccination uptake among adults is high (83%) in the study cohort, only 57% had accepted the booster dose that was generally offered to everyone by the end of follow up. We have previously reported substantially lower vaccination uptake in the study cohort among persons born outside the Nordic countries (7). In the current follow up, the overrepresentation of foreign born among cases with severe disease decreased during the transition from BA.1 to BA.2, which points at widespread dropouts from the vaccination program after two doses. This means that the overall population protection against severe COVID-19 can be insufficient in case of new intermittent epidemics that may become the “new normal” (8).

Another salient finding was that the protection associated with a prior SARS-CoV-2 infection also declined during the transition from Omicron BA.1 to BA.2. A recent total population study from Sweden reported long-lasting protection among unvaccinated persons who have survived and recovered from a previous SARS-CoV-2 infection (9). However, the follow up of that study ended before Omicron became dominant. Our study with a more recent follow up suggests that the natural protection from previous VOCs against severe disease is markedly lower when Omicron BA.2 is dominant.

The key strengths of our study were the detailed individual-level data on vaccinations, infections and hospitalisations and the possibility to stratify hospitalised people further by disease severity, thereby limiting the misclassification of cases hospitalised *with* rather than *because of* COVID-19. A major limitation was that we lacked data on virus variant for the individual cases in each time period. Our study may therefore underestimate the true change in VE associated with the transition from BA.1 to BA.2. Another limitation was that we could not evaluate protection from prior Omicron infection, as the follow up period with Omicron dominance is still short. Evidence from Qatar suggests that infection from BA.1 protects against reinfection with BA.2 but this was observed during a very short follow up period (10). A further limitation of our study was that general testing was no longer generally recommended, which means that VE against infection could not be evaluated. It should also be noted that the statistical uncertainty in some of our subgroups were substantial, as reflected in the wide confidence intervals. Continued monitoring of VE associated with Omicron BA.2 is therefore warranted.

## Conclusion

VE remained relatively stable after the transition from BA.1 to BA.2 among people with at least three doses but decreased markedly among those with only two doses. Protection from prior infection was also lower when BA.2 dominated. These findings suggest that booster vaccination is needed to maintain sufficient protection against severe COVID-19.

## Data Availability

Aggregated surveillance data from the present study are publicly available.

https://sodrasjukvardsregionen.se/kliniskastudier/covid-vacciner-skyddseffekt/

## Acknowledgments

Cecilia Åkesson-Kotsaris, Paul Söderholm and Helena Hallefjord, Clinical Studies Sweden, for excellence in bringing the surveillance infrastructure in place. Claus Bohn Christiansen, Scania Region, Clinical microbiology, for providing data from routine sequencing of infected cases.

## Authors’ contributions

JB and FK conceived the study, with important contributions from CB, MM, MR, UM and MI. UM, JB and MM acquired data, CB, JB and MM conducted the statistical analyses. JB drafted the manuscript with assistance from FK and MI. All authors contributed with interpretation of results, critically revised the manuscript and approved the final version for submission.

## References

1. Lyngse FP, Kirkeby CT, Denwood M, Christiansen LE, Mølbak K, Møller CH, et al. Transmission of SARS-CoV-2 Omicron VOC subvariants BA.1 and BA.2: Evidence from Danish Households. medRxiv. 2022.

2. Kirsebom FCM, Andrews N, Stowe J, Toffa S, Sachdeva R, Gallagher E, et al. COVID-19 Vaccine Effectiveness against the Omicron BA.2 variant in England. medRxiv. 2022.

3. Altarawneh HN, Chemaitelly H, Ayoub HH, Tang P, Hasan MR, Yassine HM, et al. Effect of prior infection, vaccination, and hybrid immunity against symptomatic BA.1 and BA.2 Omicron infections and severe COVID-19 in Qatar. medRxiv. 2022.

4. Björk J, Inghammar M, Moghaddassi M, Rasmussen M, Malmqvist U, Kahn F. High level of protection against COVID-19 after two doses of BNT162b2 vaccine in the working age population -first results from a cohort study in Southern Sweden. Infectious diseases (London, England). 2021:1–6.

5. Kahn F, Bonander C, Moghaddassi M, Rasmussen M, Malmqvist U, Inghammar M, et al. Risk of severe COVID-19 from the Delta and Omicron variants in relation to vaccination status, sex, age and comorbidities - surveillance results from southern Sweden, July 2021 to January 2022. Euro surveillance : bulletin Europeen sur les maladies transmissibles = European communicable disease bulletin. 2022;27(9).

6. Dean NE. Re: “Measurement of vaccine direct effects under the test-negative design”. American journal of epidemiology. 2019;188(4):806–10.

7. Inghammar M, Moghaddassi M, Rasmussen M, Malmqvist U, Kahn F, Björk J. COVID-19 vaccine uptake among older people in relation to sociodemographic factors – cohort results from southern Sweden. medRxiv. 2021.

8. Eales O, de Oliveira Martins L, Page AJ, Wang H, Bodinier B, Tang D, et al. The new normal? Dynamics and scale of the SARS-CoV-2 variant Omicron epidemic in England. medRxiv. 2022.

9. Nordström P, Ballin M, Nordström A. Risk of SARS-CoV-2 reinfection and COVID-19 hospitalisation in individuals with natural and hybrid immunity: a retrospective, total population cohort study in Sweden. The Lancet Infectious diseases. 2022.

10. Chemaitelly H, Ayoub HH, Coyle P, Tang P, Yassine HM, Al-Khatib HA, et al. Protection of Omicron sub-lineage infection against reinfection with another Omicron sub-lineage. medRxiv. 2022.

