## Supplementary Material for "COVID-19 vaccine effectiveness against severe disease from the Omicron BA.1 and BA.2 subvariants – surveillance results from southern Sweden, December 2021 to March 2022"

**Content**

Supplementary Table S1
Supplementary Table S2
Supplementary Table S3

**Supplementary Table S1.** Routine sequencing of samples of infected cases in Scania county, Sweden.

| Year | 2021 | 2022 | | | | | | | | | | |
|---|---|---|---|---|---|---|---|---|---|---|---|---|
| Week | 52 | 1 | 2 | 3 | 4 | 5 | 6 | 7 | 8 | 9 | 10 | 11 |
| Total | 289 | 325 | 281 | 340 | 288 | 314 | 314 | 222 | 222 | 123 | 124 | 157 |
| Omicron BA.1 | 162 | 206 | 136 | 153 | 72 | 65 | 40 | 32 | 77 | 6 | 6 | 6 |
| Omicron BA.2 | 29 | 61 | 124 | 179 | 214 | 244 | 273 | 190 | 144 | 117 | 118 | 151 |
| Delta | 98 | 58 | 21 | 8 | 2 | 5 | 1 | 0 | 1 | 0 | 0 | 0 |

**Supplementary Table S2.** Classification of comorbidities.

| Disease group | ICD-10 codes (incl KVÅ-codes[a]) |
|---|---|
| Cardiovascular diseases | I10-I15, I20-I25, I42-I43<br>I50, I60-I69<br>J81 |
| Diabetes or obesity | E10, E11, E66 |
| Kidney or liver diseases | K70.X, K74.3-K74.6, K75.4, K76.0<br>N18.5, N18.9<br>DR016, DR024 |
| Respiratory diseases | A15-A19<br>E84<br>I26, I27<br>J42, J43, J44, J45, J47, J84<br>J96, J98.2, J98.3 |
| Neurological diseases (including dementia) | G00-G99<br>F00-F03 |
| Cancer or immunosuppressed state (including organ transplantation) | C00-C99<br>KAS, FQA, FQB, JJC, GDG, JLE<br>DR046, DR047, DR048<br>D80.0-D80.1<br>D80.5, D81, D82, D83 |
| Other conditions and diseases<br>• HIV<br>• Thalassemia<br>• Sickle cell<br>• Mood disorders<br>• Schizophrenia spectrum disorders<br>• Substance use disorders<br>• Downs syndrome | B20-B24<br>D56, D57<br>F10-F19, F30-F39, F20-F29<br>Q90 |

[a] Swedish classification of certain interventions during health care visits

**Supplementary Table S3.** Vaccine effectiveness after at least three or two doses against severe COVID-19 in each follow up period. Estimates were obtained from conditional logistic regression for age and sex matched case and controls (1:10), and with adjustment for comorbidities (0, 1, ≥2) and infection at least 90 days prior the case date. Results are presented overall and stratified by age, sex and comorbidities.

| | Follow up period | | |
|---|---|---|---|
| | Omicron BA.1<br>2021 w52-2022 w1 | Transition<br>2022 w2-3 | Omicron BA.2<br>2022 w4-11 |
| | VE (95% CI)[a] | VE (95% CI)[a] | VE (95% CI)[a] |
| *At least three doses* | | | |
| All | 94 (84 – 98) | 90 (82 – 95) | 82 (64 – 91) |
| Age | | | |
| < 65 years | 92 (73 – 98) | 74 (16 – 92) | 76 (32 – 91) |
| ≥ 65 years | 94 (76 – 98) | 92 (83 – 97) | 82 (56 – 93) |
| Sex | | | |
| Females | 92 (74 – 97) | 93 (81 – 98) | 78 (35 – 93) |
| Males | 96 (79 – 100) | 89 (74 – 96) | 84 (62 – 93) |
| Comorbidities | | | |
| None | 99 (53 – 100) | 96 (81 – 100) | 77 (27 – 92) |
| ≥ 1 | 90 (44 – 98) | 84 (54 – 95) | 85 (48 – 96) |
| *Two doses* | | | |
| All | 90 (78 – 95) | 70 (42 – 84) | 54 (13 – 75) |
| Age | | | |
| < 65 years | 93 (81 – 98) | 78 (35 – 93) | 59 (4 – 83) |
| ≥ 65 years | 84 (37 – 96) | 55 (0 – 81) | 43 (0 – 79) |
| Sex | | | |
| Females | 88 (65 – 96) | 73 (37 – 88) | 53 (0 – 82) |
| Males | 92 (73 – 98) | 67 (14 – 87) | 54 (0 – 80) |
| Comorbidities | | | |
| None | 96 (66 – 100) | 79 (32 – 94) | 61 (2 – 85) |
| ≥ 1 | 77 (0 – 95) | 58 (0 – 86) | 46 (0 – 86) |

[a]Vaccine effectiveness (95% confidence interval)
